# Knowledge, Attitude, and Practices towards SARS-COV-2 Infection in the United Arab Emirates Population: a Cross-sectional Survey-based Study

**DOI:** 10.1101/2021.02.24.21252331

**Authors:** Hamda Musabbah Alremeithi, Aljazia Khalfan Alghefli, Rouqyah Almadhani, Latifa Baynouna AlKetbi

## Abstract

In the current situation involving the novel coronavirus disease (COVID-19), the disease containment depends mainly on the population’s preventive practices and self-isolation. In this study, we explored the adult population’s approach towards COVID-19 in UAE between the 4^th^ and 14^th^ of April 2020. We used a community-based, cross-sectional study using a self-administered electronic questionnaire covering five different aspects: demographics, knowledge, practice, attitude, source and trust of information, and a patient health questionnaire (PHQ-2) for depression screening. A total of 1,867 people responded to the survey. Knowledge results were better in people with higher education levels, non-UAE nationals, those with a higher PHQ-2 score, or with a positive contact history with a SARS-COV-2 infected patient. The best practice scores were shown by participants with high knowledge scores and education levels. Depression risk was significantly higher in men, non-UAE nationals, in those with lower knowledge scores, and younger ages. The most followed practices were staying home, handwashing, avoiding social gatherings, limiting three people per vehicle, and avoiding public transportation. The least practiced measures were covering the face while sneezing or coughing and wearing masks. Although staying home was reported by 92.5% of participants, 22.6% mentioned that they were visited by more than 2 people and visited others in 18.4% during the last week. Social media was the source of information for 82.1% of the participants, and most trusted doctors and healthcare providers. A depression risk was present in 18.9% of the participants and the majority of respondents agreed that SARS-COV-2 infection will finally be successfully controlled. The obtained results on knowledge and practices, although satisfactory, could be insufficient to prevent this pandemic from being contained. We recommend the intensification of awareness programs and good practices. Mental health is an area worth further studies.

## Introduction

The emerging pandemic that started at the end of 2019 in Wuhan, China, has tremendously affected the life of the global population. No event in recent history had such widespread impact. The SARS-COV-2 infection is an infectious disease caused by a new coronavirus that causes a wide variety of symptoms. Although most people infected with SARS-COV-2 have mild to moderate disease, people who have underlying comorbidities might develop a more serious life-threatening illness; moreover, due to its high infectivity, the gravely affected patients have crippled healthcare systems across the world. The disease was contained in the city of its origin mainly through population mass isolation and social distancing, with strict strategies to limit the infection rate among the population (1).

The world health organization (WHO) and the United States Center for Disease Control and Prevention (CDC) released recommendations to control the spread of the infection, which were rapidly implemented by many governments with variable success rates. The focus was on improving the population’s infection control measures. These measures were extended to total lockout in some countries, with a stay-at-home directive, closed schools, and cancelled sporting and cultural events (2). To date, although no specific vaccines or treatments for the novel coronavirus disease (COVID-19) have been found, many ongoing clinical trials have been evaluating the potential treatments (1).

Despite an increasing incidence of COVID-19 in the United Arab Emirates, to date no study has assessed the knowledge, attitude, and practices of local communities regarding COVID-19 transmission and its prevention. Containment of this infection is mainly dependent on the population’s preventive practices and self-isolation, and its high transmissibility is caused by a high level of SARS-COV-2 shedding in the upper respiratory tract, even among presymptomatic patients who may spread the infection without knowing (1) (3) (4). Therefore, studying the knowledge, attitude, and practices of the population regarding virus transmission is a determinant step in the containment efforts of the pandemic. The aim of the present study was to assess these aspects in the United Arab Emirates and identify its determinants to design intervention strategies for an effective prevention program.

## Materials and Methods

### Study design

This community-based cross-sectional survey assessed the knowledge, attitude, and practices towards infection by SARS-COV-2.

### Target population and sampling procedure

Data was collected during 4^th^-14^th^ April 2020 over a period of ten days. In total, 1992 people responded to the survey, 1867 of which were included in the analysis as the others were from outside the UAE. Assuming a precision of 5% and a 95% confidence level, the minimum required sample size should have been 1056, and our number was in accordance to this requirement. Convenient sampling was possible owing to the use of an electronic survey distributed through social media (WhatsApp, Instagram, and Snapchat applications). The following inclusion criteria were applied: people over 16 years old and living in the United Arab Emirates.

### Study instrument

At the time of the study design, there was no validated questionnaire for SARS-COV-2 infection; therefore, a new questionnaire was designed and structured based on previous studies assessing knowledge, attitude, and practices towards previous outbreaks (5, 6). The questionnaire was further adjusted to accommodate the emerging SARS-COV-2 infection using available data obtained in a literature review(7).The questionnaire was constructed to cover five main domains: 1. demographic data; 2. knowledge assessment; 3. practice assessment; 4. attitude assessment (Appendix 1); and 5. patient health questionnaire-2 (PHQ-2) as a screening tool for depression among participants.

To measure the knowledge towards SARS-COV-2 infection, we used eight multiple-choice questions with answers including the “I don’t know” option and one open-ended question about safe social distancing. Questions developed using the information obtained in the literature review were used to measure knowledge regarding the causative agent, transmission, symptoms, high risk groups, and prevention and precaution measures. The accumulative score was the resultant of adding all correct answers. Higher scores thus indicated better knowledge on these aspects.

A set of 10 questions adapted from previous studies was used to measure the preventive practices. Participants were asked to respond to these questions on a five-point Likert-like scale indicating the frequency of practicing the presented statements, ranging from “always” to “never”. The assessment of participants’ practices evaluated the following behaviors: handwashing, use of hand sanitizer and face masks, keeping a safe social distance, covering mouth and nose during sneezing and coughing, using the non-dominant hand whenever in public, avoiding face touching and going out, avoiding the use of public transportation, limiting up to 3 people per vehicle, and avoiding family gatherings and social events.

We elaborated more on panic shopping by asking 2 questions: 1-have you done extra grocery shopping beyond your personal needs after hearing about the spread of SARS-COV-2? 2-How long do you have enough groceries for? Focusing on social contacts made in the previous week, we asked the following: how many times did you go out during the last week? How many times did you visit someone at their home during the last week? How many people visited you last week? How many delivery or maintenance workers visited your home during the last week? Finally, the attitude towards SARS-COV-2 infection was measured by asking if the participant was willing to take the vaccine when available. Information sources and the participants’ trust in their information resources were assessed.

### Risk assessment

Personal history of recent travel or contact with COVID-19 cases were investigated as these aspects could be the determinants of the study aim. General health and comorbidities were also investigated as they can influence individual knowledge, attitude, and practices. The questionnaire included questions on conditions like pregnancy, cardiovascular disease, hypertension, diabetes mellitus, cancer, and chemotherapy and use of immunosuppressants. According to the risk assessment questions, we classified respondents in low-risk and high-risk groups (Appendix 2).

The resultant questionnaire was pilot tested for content and face validity among 30 participants, and minor revisions were made. The final questionnaire, which was in English, was translated to Arabic and both questionnaire, Arabic and English versions,were distributed to the community by a Survey Monkey (www.surveymonkey.com) link sent through social media. Participation was completely voluntary. This questionnaire distribution method was found suitable for reaching large numbers of people in the community, and also for avoiding close personal contact and preventing COVID-19.

### Statistical analysis

This research was approved by the Ambulatory Healthcare Services’ Human Ethics Committee and the Abu Dhabi Healthcare Services Central Human Ethics Committee. Statistical analysis was performed using SPSS StatisticsVersion 23(IBM, Armonk, USA). Results were analyzed using frequencies, cross-tabulation, and regression analysis.

## Results

The study had an excellent response rate with most of the samples collected within 3 days. We obtained a sample of 1882 people, considering 1904 who completed the survey and those excluded for being from outside the UAE and under 16 years of age. In the final sample, most respondents were female (80.7%), from all age groups: 67.7% were younger than 40 and 32.3% were older than 40 years old. UAE nationals were the majority of respondents (1471, 78.2%) and most of them were from Abu Dhabi (1179, 63.6%). Government employees were 814 (44.8%), and 1604 (87.7%) were working from home, students, or unemployed at the time of the questionnaire. Approximately half the respondents (1018, 54.5%) held a bachelor’s degree or above. Three quarters of respondents were healthy (1380, 73.3%), with no underlying medical conditions (Table 1).

**Table 1.**
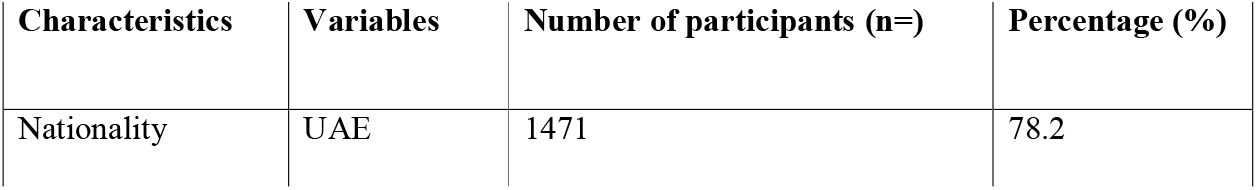
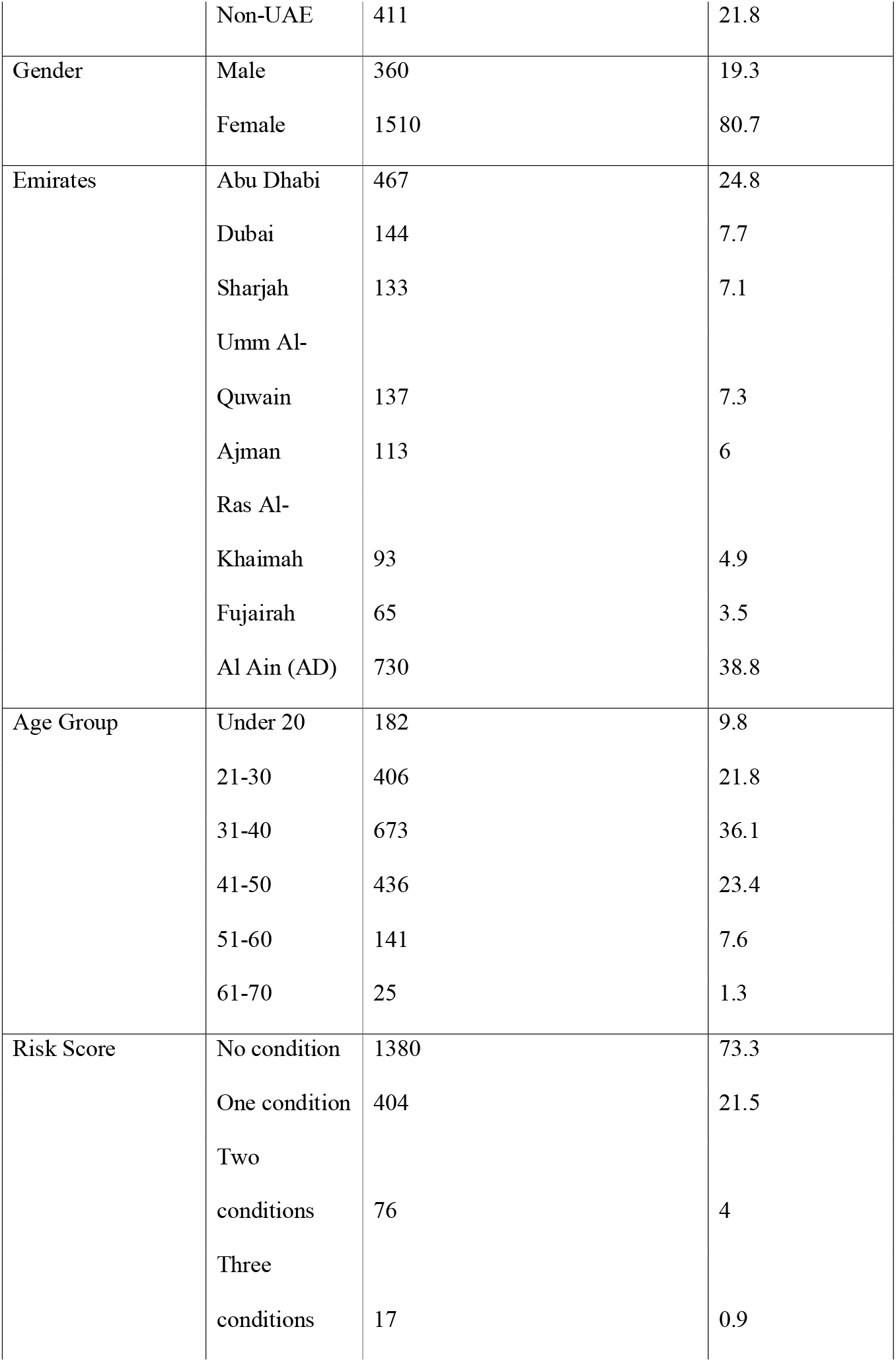
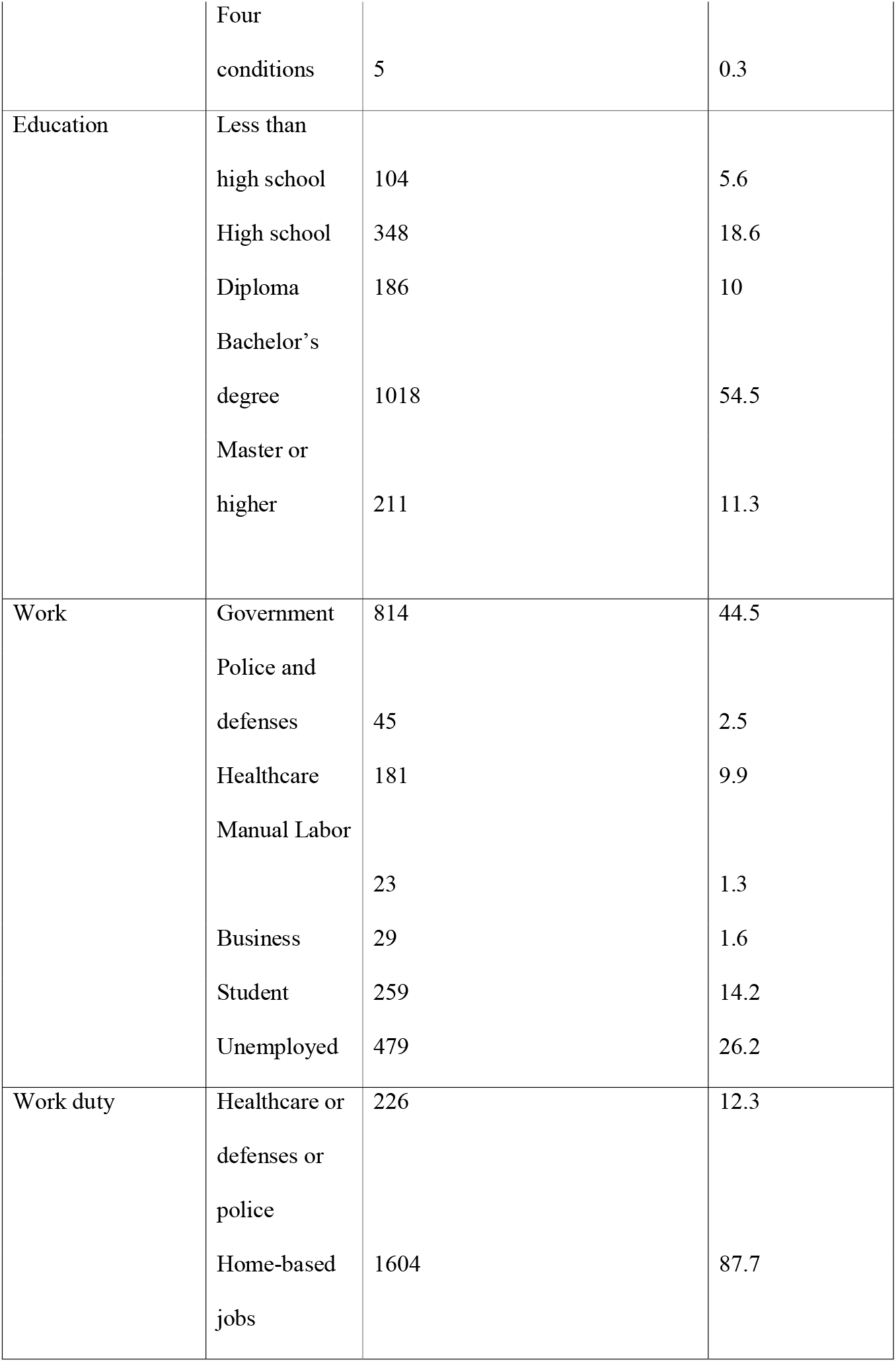
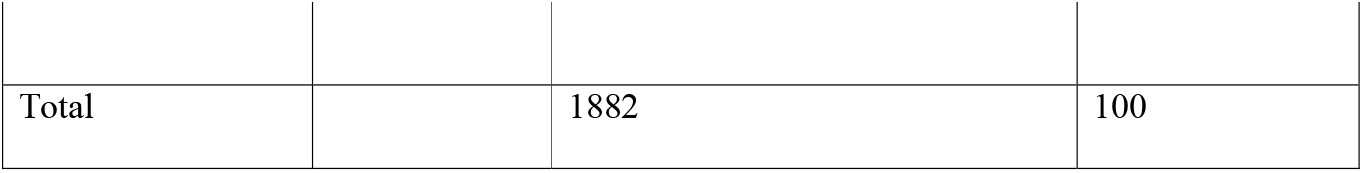
Demographic characteristics of participants and knowledge score by demographic variables.

| Characteristics | Variables | Number of participants (n=) | Percentage (%) |
| --- | --- | --- | --- |
| Nationality | UAE | 1471 | 78.2 |
|  | Non-UAE | 411 | 21.8 |
| Gender | Male | 360 | 19.3 |
|  | Female | 1510 | 80.7 |
| Emirates | Abu Dhabi | 467 | 24.8 |
|  | Dubai | 144 | 7.7 |
|  | Sharjah | 133 | 7.1 |
|  | Umm Al- |  |  |
|  | Quwain | 137 | 7.3 |
|  | Ajman | 113 | 6 |
|  | Ras Al- |  |  |
|  | Khaimah | 93 | 4.9 |
|  | Fujairah | 65 | 3.5 |
|  | Al Ain (AD) | 730 | 38.8 |
| Age Group | Under 20 | 182 | 9.8 |
|  | 21-30 | 406 | 21.8 |
|  | 31-40 | 673 | 36.1 |
|  | 41-50 | 436 | 23.4 |
|  | 51-60 | 141 | 7.6 |
|  | 61-70 | 25 | 1.3 |
| Risk Score | No condition | 1380 | 73.3 |
|  | One condition | 404 | 21.5 |
|  | Two |  |  |
|  | conditions | 76 | 4 |
|  | Three |  |  |
|  | conditions | 17 | 0.9 |
|  | Four conditions | 5 | 0.3 |
| Education | Less than high school | 104 | 5.6 |
|  | High school | 348 | 18.6 |
|  | Diploma | 186 | 10 |
|  | Bachelor's degree | 1018 | 54.5 |
|  | Master or higher | 211 | 11.3 |
| Work | Government | 814 | 44.5 |
|  | Police and defenses | 45 | 2.5 |
|  | Healthcare | 181 | 9.9 |
|  | Manual Labor | 23 | 1.3 |
|  | Business | 29 | 1.6 |
|  | Student | 259 | 14.2 |
|  | Unemployed | 479 | 26.2 |
| Work duty | Healthcare or defenses or police | 226 | 12.3 |
|  | Home-based jobs | 1604 | 87.7 |
| Total |  | 1882 | 100 |

## Knowledge

Among all respondents, 79.6% answered 76.5% of the questions correctly (13/17). The mean SARS-COV-2 infection knowledge score was 11.5 (standard deviation, SD 2.5), and only 18.3% scored less than 10. The knowledge score was significantly determined by higher educational levels (B 0.18, *P* value < 0.05), good preventive practice (B 0.121, *P* value < 0.05), and higher risk scores (B 0.053, *P* value < 0.05) (Table 2). Interestingly, only half (52.2%) of the healthcare providers correctly answered the recommended social distance of 2 meters; this result does not differ much from the other participants (47%).

**Table 2.**
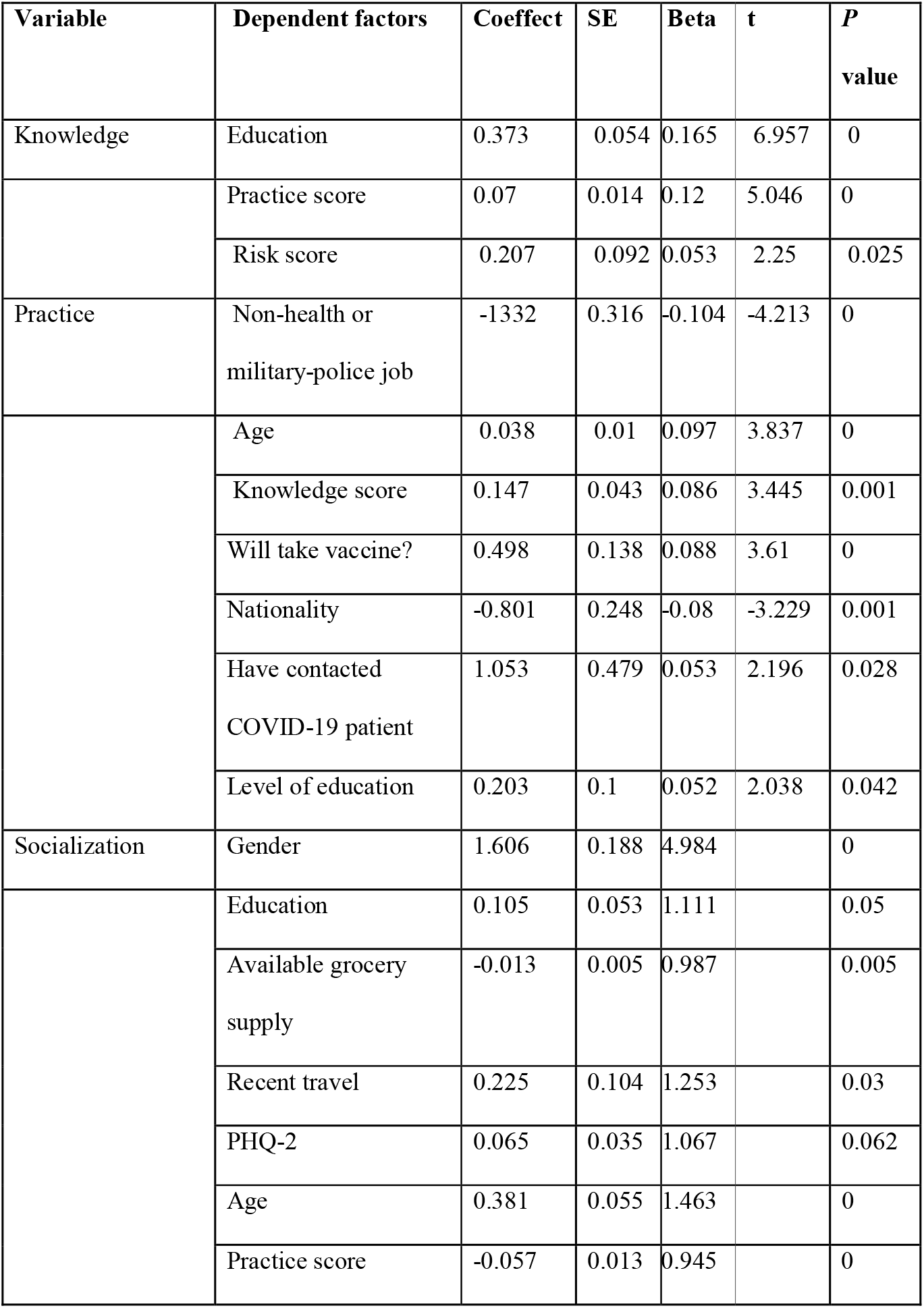
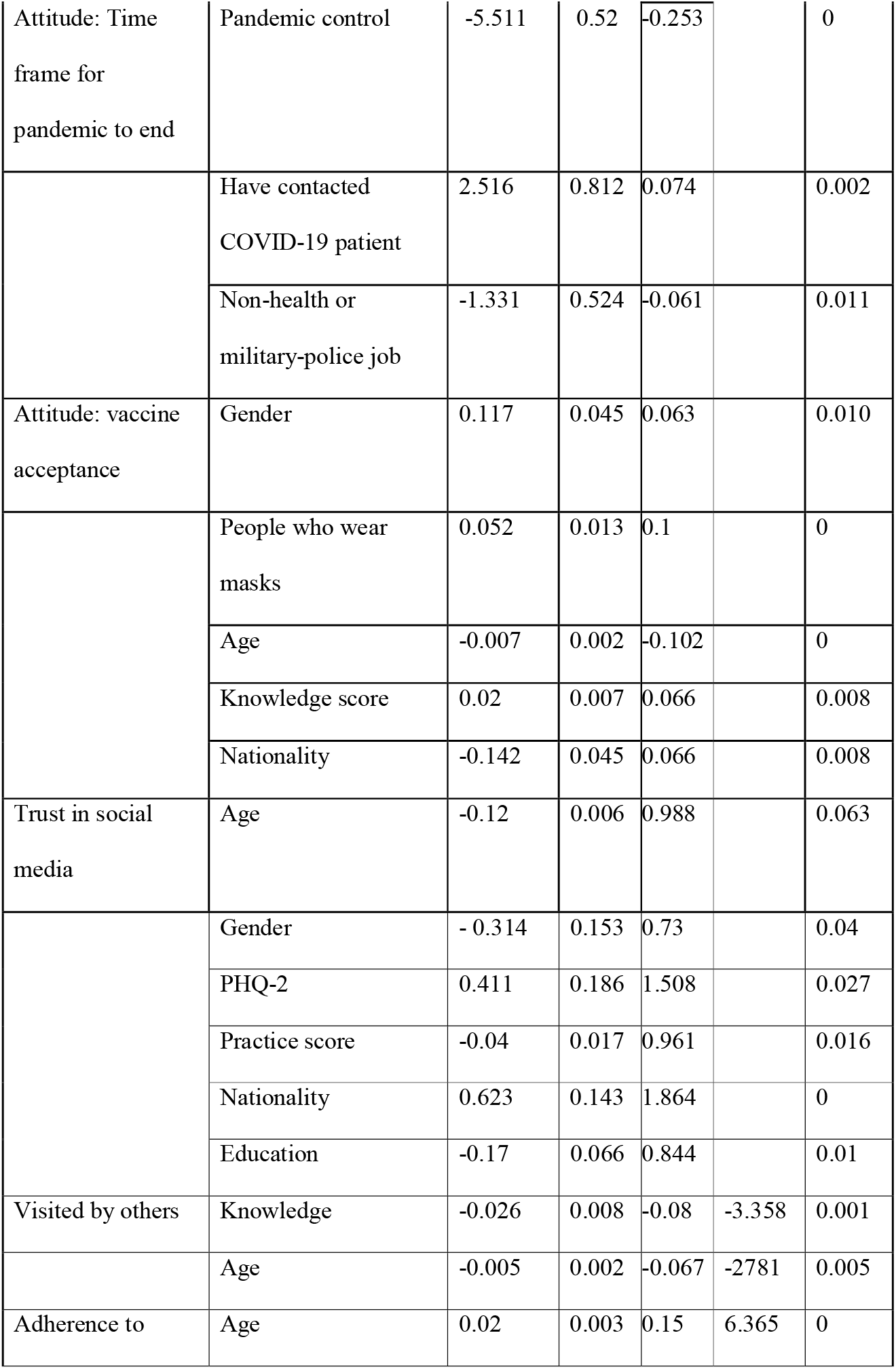
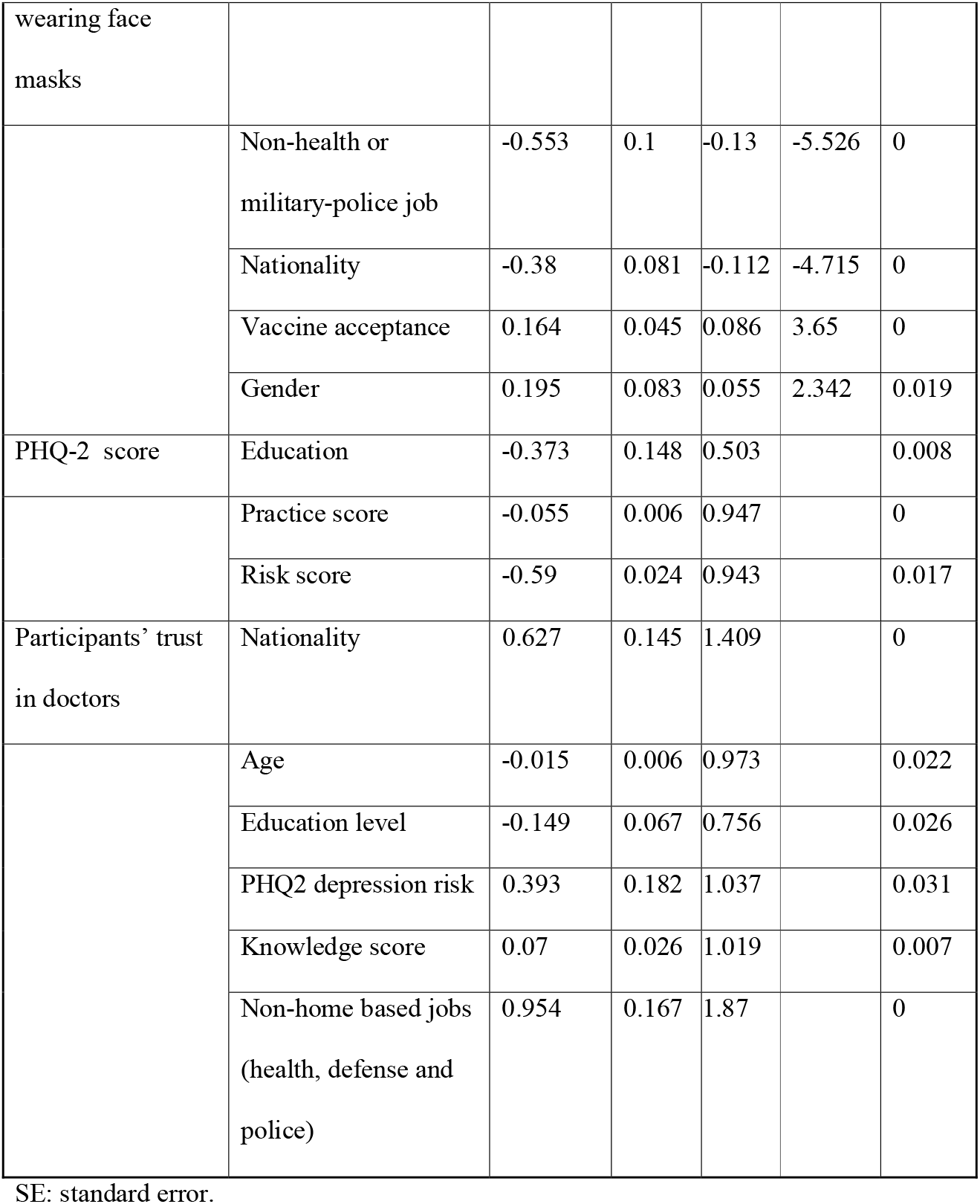
Regression determinant factors.

### Practice

The survey uncovered important determinants of the population with regards to choices and adherence to safe practices in the pandemic. Although excellent adherence was reported, with more than 90% reporting staying home, 28.5% and 6.2% reported having visited or been visited by someone. In addition, 59.4% received a delivery worker in their homes at least once. Two thirds of the respondents achieved a score of 78.9% (31 points out of 38) on the best practices in preventing SARS-COV-2 infection. The mean score on SARS-COV-2 infection prevention practices was 29.5 (SD 4.2). Respondents who were more likely to adhere to better practices were those of older age (B 0.097, *P* value < 0.05), with good knowledge (B 0.086, *P* value < 0.05), were of non-UAE nationalities (B −0.08, *P* value < 0.05), with jobs that cannot be practiced from home, military and health care employees (B - 0.104, *P* value < 0.05), had a personal history of contact with COVID-19 patients (B 0.053, *P* value < 0.05), higher educational levels (B 0.052, *P* value < 0.05), and a positive attitude towards taking a vaccine (B 0.088, *P* value < 0.05) (Table 2).

People who were more likely to socialize and not stay at home during the pandemic were male (B 1.6, *P* < 0.05), with higher education levels (B 0.105, *P* < 0.05), had a smaller grocery supply (B −0.013, *P* < 0.05), had personal history of recent travel (B 0.225, *P* < 0.05), scored higher in the PHQ-2 questionnaire (B 0.065, *P* < 0.05), were older in age (B 0.381, *P*<0.05), and presented poor scores on prevention practices (B −0.057, *P* < 0.05). Results showed that people who were more likely to visit others were of younger age (B −0.067, *P* < 0.05) and had inferior knowledge on the pandemic (B −0.086, *P* < 0.05) (Table 2)

Wearing face masks in public is considered one of the most important measures to prevent transmission of this infection (8). Nevertheless, only one third (32.2%) of the participants reported always wearing a mask (Figure 1). People who were compliant to wearing masks were more likely to be older in age (B 0.15, *P* < 0.05), work in health or military fields (B 0.13, *P* < 0.05), be non-UAE nationals (B −0.112, *P* < 0.05), have a positive attitude towards taking a vaccine (B −0.086, *P* < 0.05), and be male (B 0.055, *P* < 0.05) (Table 2).

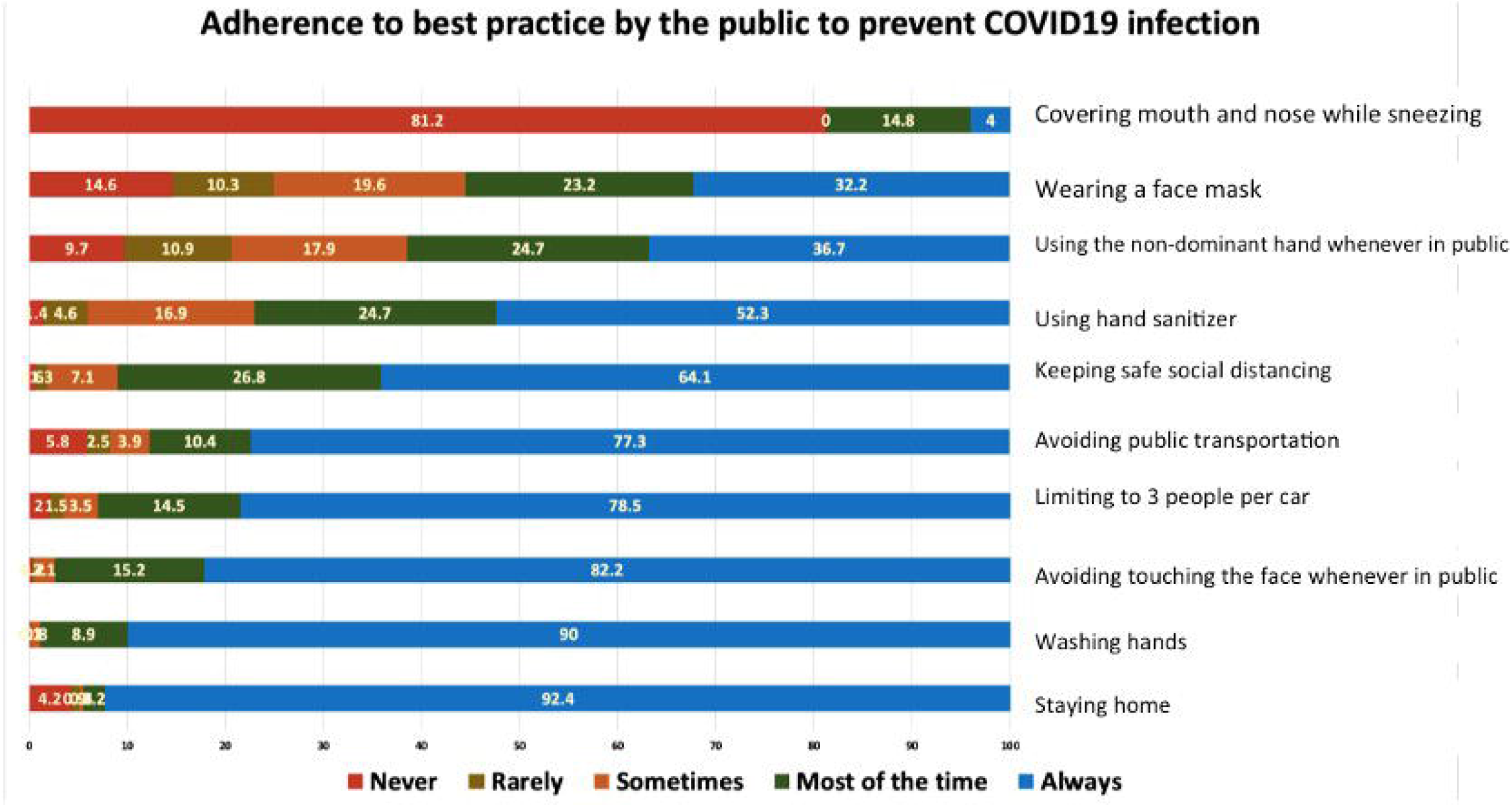

Among the participants, 181 (9.9%) were healthcare workers. It is important to highlight their practices, where 69% reported always keeping social distancing, 44.8% always wearing a face mask, 93.9% always washing hands, and 73.5% always using hand sanitizer.

### Attitude

The majority of the respondents (89%) agreed that COVID-19 will finally be successfully controlled; 37.5% believed that this pandemic would end within 2-3 months (Table 2). Participants’ beliefs in optimistic time frames to control the pandemic depended on the nature of their work area: more optimism was perceived among healthcare workers and military (B - 0.061, *P* value < 0.05) and less optimism was shown by those with personal history of contact with COVID-19 patients (B-0.074, value<0.05).

People who were more likely to accept a vaccine wore masks (B 0.01, *P* value < 0.05), were of younger age (B −0.102, *P* value < 0.05), non-UAE nationals (B −0.08, *P* value < 0.05), had higher knowledge scores (B 0.066, *P* value < 0.05), and were male (B 0.063, *P* value < 0.05) (Table 2).

### Trust and sources of information

Most of the participants trusted doctors and healthcare providers as sources of information (56.7%). These participants were younger in age (B −0.015, *P* < 0.05), UAE nationals (B 0.627, *P* < 0.05), with lower educational levels (B −0.149, *P* < 0.067), had good knowledge scores (B 0.07, *P* < 0.05), scored higher in risk of depression (B 0.393, *P* < 0.05), and worked in non-homebased jobs such as healthcare, police, or army forces (B 0.954, *P* < 0.05).

Regarding the respondents’ sources of information, social media was the source of information of 1545 (82.1%) participants. Television news came next as a source of information for 1001 participants (53.2%). Those using social media as a source of information were more likely to be UAE nationals (B 0.627, *P* < 0.05), younger in age (B − 0.015, *P* < 0.05), with a lower educational level (B 0.149, *P* < 0.05) and a higher risk of depression (B 0.393, *P* < 0.05), with high knowledge scores (B 0.07, *P* < 0.05), and were more likely to be military or health care employees (B 0.954, *P* < 0.05).

### Mental health during the pandemic

Almost one in five (18.9%) participants presented a depression risk, which was assessed using a PHQ-2 questionnaire (Appendix 3). This value represents twice what was reported last year among ambulatory health services patients (10%) (8). We observed that depression risks were significantly associated with lower knowledge scores (B −0.059, *P* < 0.05), younger age (B −0.055, *P* < 0.05), and non-UAE nationality (B −0.0395, *P* < 0.05) (Table 2).

## Discussion

To the best of our knowledge, this is the first study in the UAE and Eastern Mediterranean region to assess the knowledge, attitude, and practices towards SARS-COV-2 infection. Almost 80% of the respondents answered correctly to 76% of the knowledge questions, indicating that most respondents were knowledgeable about SARS-COV-2 infection. However, there are still areas of concern, such as social distancing (half of the respondents did not know the correct safe distance between people). Since this study was conducted during the early to intermediate stage of the COVID-19 pandemic, we expected good knowledge and practice results towards SARS-COV-2 infection among UAE residents.

The selected population had good knowledge scores due to the sample characteristics, where 75.8% held a diploma degree or higher. Our study also showed a significant positive association between the level of knowledge and a participant’s risk score, where assessment of the risk depended on the presence of medical conditions that lower the participant’s immunity, making them more susceptible to being infected (2). This study participants would actively learn about the infection from trusted resources. The significant positive association between level of education and SARS-COV-2 infection knowledge score supports this speculation. A Chinese study has recently found a similar high knowledge rate among its participants (90%), indicating that the Chinese population is knowledgeable about SARS-COV-2 (9). However, according to Haque *et al*., among 2045 respondents across Bangladesh, only 54.87% had good knowledge regarding COVID-19. This study showed that knowledge on SARS-COV-2 infection among Bangladeshi people depended significantly on age, gender, education level, place of residence, income, and marital status (10).

Regarding practices, the majority of participants reported staying at home (92.8%), which was expected considering the national awareness programs and the adherence to the sanitization campaigns across the Emirates. The same applies to aspects as washing hands and avoiding touching one’s own face. Although high knowledge was associated with better preventive practices, the adherence to some practices such as covering the face while sneezing and coughing (never = 81.2%) and wearing a mask (always or sometimes = 55.4%) was suboptimal. The most concerning result is probably the lack of face mask use. When evaluating the current literature, we observed that although there are no available randomized controlled trials (RCTs) that prove the effectiveness of mask wearing on preventing SARS-COV-2 transmission, studies report that a critical practice to reduce transmission of a respiratory viral infection is the use of face masks, surgical masks, or N95 respirators (10).Initial Chinese preventive recommendations and public practice guidelines highlighted the use of face masks for protection against SARS-COV-2 (11).These recommendations are reflected by the results obtained by Zhong *et al*. (2020), where 98% of the respondents in China reported wearing a face mask when going out (9). A study performed in India showed that only 76% of the respondents used face masks (9). Considering the UAE population, we highlight the need for a stronger adherence to the prevention measures in order to control the current pandemic.

In the absence of available vaccines and proven treatments, the only available public health tools to control person-to-person transmission are isolation and quarantine, social distancing, and community containment measures(5). Our study results show the lack of adherence to home quarantine, since 81.6% of the respondents visited someone and 77.4% were visited in the previous week. Arab culture and extended family relations in the region can explain the poor adherence to the practices related to social relations. When comparing our results to those obtained in China, Zhong *et al*. (2020) reported better practices, where 96.4% of the participants had not visited crowded places and 98% wore masks when going out in recent days(12). More adhereance to wearing mask in china, Zhonge et al (2020) might be attrubitted to the fact that particepent were visting public crowded places, and less adherenace to wearing mask in our study might be related to the fact that paritecepts were visting or being visted by family members.

The rationale behind social distancing is to slow or stop the spread of COVID-19, which requires full engagement of all members of society and the cancellation of social events and gatherings (2). The most effective public health measures observed in China were community isolation with social distancing, community use of facemasks, and a lockdown of Wuhan’s public transportation, including buses, trains, ferries, and the airport (13). This was the biggest quarantine in history, starting on January,23,2020 and ending on April,8,2020 and lasted for a total of 76 days., where 113,579 close contacts were tracked and 102,427 were under medical observation. This approach was a remarkable effort that was effective in the successful SARS-COV-2 containment.

Most of the participants of our study (70.67%) held an optimistic attitude towards the COVID-19 pandemic: 37.50% believed that it would finally be successfully controlled within 2-3 months, while 33.17% believed that it would be controlled in less than 2 months. These findings can be explained by the trust in government authorities, the local healthcare system, and the country’s infrastructure by UAE residents. People who believed that infection control would take longer tended to be non-healthcare workers, military, police officers, and those who had contact with people suffering from SARS-COV-2 infection. More optimistic findings were observed among Chinese people, where 90.8% believed that COVID-19 would finally be successfully controlled, and 97.1% were confident that China could control the virus(12).

Identifying determinants of better knowledge and practices such as higher education levels, higher risk scores, non-local nationality, previous contact with a COVID-19 patient, older age, and healthcare and military workers, is useful for public health policy makers and health workers to recognize the target population for education on health and disease prevention. In participants with lower educational levels, improving health literacy is a determinant factor in the pandemic containment. At the time of our survey, the UAE government instituted social isolation from 8 pm to 6 am, closing schools and universities, instructing people to work from home when applicable, and cancelling all social events. Nevertheless, the adherence of the population, as reported in this study, is considered satisfactory but not optimal to prevent the transmission of the disease.

The relationship between depression risk and the SARS-COV-2 pandemic was assessed only in one study to date. In a population-based study, being female and a student, as well as having symptoms suggestive of COVID-19 and poor perceived health were associated with higher rates of anxiety and depression; on the other hand, the availability of accurate information and the use of specific preventive measures, such as handwashing, seemed to mitigate these effects(9). In our study, better knowledge was also protective against depression risk, highlighting an aspect of depression prevention that could be focused by strong awareness programs, especially among the young who showed a higher depression risk in this study.

Unexpectedly, the practices of UAE healthcare workers as assessed by our study were suboptimal. A study performed in Nigeria reported low knowledge (78.6%) and poor attitude (64%) among healthcare workers (Ayinde et al., 2020). On the contrary, Saqlain *et al*.’s findings showed that Pakistani healthcare workers had good knowledge (93.2%, n = 386), positive attitude (8.43 ± 1.78), and good practices (88.7%, n = 367) regarding SARS-COV-2 (14). Similar findings by Giao *et al*. illustrated that the majority of participants among healthcare workers in Vietnam held good knowledge and good attitude towards SARS-COV-2 (15). More research is needed to investigate healthcare workers’ competences and the knowledge necessary for their work in this challenging pandemic environment.

The strength of this study relies in its large sample size, recruited during a short period of time in the middle stage of the SARS-COV-2 pandemic in the UAE. The UAE is one of the countries with the highest access to internet and use of social media,thus an online survey is able to reach wide sections of the society. Nevertheless, when comparing our results to the most recent national population statistics of the UAE, our sample was obviously over-representative of women, well-educated people, and young adults. Age of the particepnts might be affected by the mean by which the questionnaire was distributed since most of social media users are young adults. Given the significant association revealed by this study between these demographic variables and the knowledge, attitude, and practices towards COVID-19, we may have overestimated the knowledge and preventive practices in the UAE population.

We spaculate that the presence of positive attitude towards COVID-19 may also differ depending on socioeconomical status. Vulnerable populations such as the elderly and manual labor workers could have been underestimated in our sample. Another limitation is that the survey was only translated to Arabic and English, underestimating the participation of speakers of other languages. Additionally, a limitation of our study is the development of an objective assessment tool of practices towards SARS-COV-2 infection. Due to the very limited time for developing our questionnaire, we measured practices by simple self-reported questions only.

The UAE residents with higher education levels, good practices, and higher risk scores have good knowledge about SARS-COV-2 infection and satisfactory practices and attitude, which suggests that health education programs should aim to improve the population’s adherence to the safe practices. With the size and impact of this pandemic, lack of adherence even by a minority could be enough to prevent the pandemic from being contained. The recommendations obtained from this study are the intensification of awareness programs and interventions, and an increase in good practices. Moreover, the COVID-19 pandemic has negatively affected the community’s mental health, therefore more attention should be dispensed on the psychiatric impact of this condition and healthcare providers should consider tackling this issue when evaluating their patients.

## Data Availability

Available on requested

## Acknowledgments

The authors thank all participants involved in this study for their cooperation and support.

The authors declare that there is no conflict of interest.

## Appendix 1. Knowledge, attitude, and practices towards SARS-COV-2 among UAE residents: questionnaire

**Appendix 1. Knowledge, attitude, and practices towards SARS-COV-2 among UAE residents: questionnaire:**
| **Questionnaire of knowledge towards SARS-COV-2 INFECTION:** |  |
| --- | --- |
| Questions | Answer options |
| K1. Corona (SARS-COV-2 INFECTION) is considered: | Bacteria, gas, virus, fungus, mosquito bite, unknown, I don't know |
| K2. A person may have Corona (SARS-COV-2 INFECTION) if he/she has the following (choose all that apply): | Fever, cough, sneezing, no symptoms, I don't know |
| K3. Corona (SARS-COV-2 INFECTION) virus is spread by (Choose all that apply): | Air, droplets, touching hard surfaces, food, I don't know |
| K4. The following are effective measures to decrease the possibility of getting Corona (SARS-COV-2 INFECTION)? (Choose all that apply) | Drinking extra water, hand washing, using hand sanitizer, wearing a mask, taking a multivitamin supplement, I don't know |
| K5. How many of those infected with Corona (SARS-COV-2 INFECTION) will recover? | One third of them, half of them, more than 90% of them, I don't know |
| K6. It is not necessary for children and young adults to adopt measures to prevent Corona (SARS-COV-2 INFECTION) infection as they are not at high risk of death or complications? | True, false, I don't know |
| K7. If isolation is required, it should be for how long? | One week, two weeks, three weeks, four weeks, I don't know |
| K8. People returning from travel should avoid going out in public and adhere to home quarantine for at | One week, two weeks, three weeks, four weeks, I don't know |
| least how long? |  |
| K9. How many meters are considered a safe social distance between you and others? | (No option, one answer) |
| **Questionnaire of practices towards SARS-COV-2 INFECTION:** |  |
| **Questions** | **Answer options** |
| P1. During last week, how often did you wash your hands? | Always, most of the time, sometimes, rarely, never |
| P2. During last week, how often did you use hand sanitizer? | Always, most of the time, sometimes, rarely, never |
| P3. During last week, how often did you use a face mask? | Always, most of the time, sometimes, rarely, never |
| P4. During last week, how often did you keep social distancing? | Always, most of the time, sometimes, rarely, never |
| P5. During last week, how often did you cover your mouth and nose if sneezing or coughing? | Always, most of the time, sometimes, rarely, never |
| P6. During the last week, how often did you use your non-dominant hand whenever in public? | Always, most of the time, sometimes, rarely, never |
| P7. Did you avoid touching your face (mouth, nose, and eye) whenever in public? | Always, most of the time, sometimes, rarely, never |
| P8. During the last week how often did you stay home and avoid going out unless for necessary purposes? | Always, most of the time, sometimes, rarely, never |
| P9. During the last week how often did you avoid using public transportation and being in crowded places? | Always, most of the time, sometimes, rarely, never |
| P10. During the last week how often did you have 3 | Always, most of the time, sometimes, rarely, |
| people or less per vehicle during transportation? | never |
| P11. During the last week how often did you avoid family gatherings and other social events? | Always, most of the time, sometimes, rarely, never |
| P12. Have you done extra grocery shopping beyond your needs after hearing about the spread of the Corona (SARS-COV-2 INFECTION) infection? | ☐Yes ☐No ☐Sometimes |
| P13. For how long do you have enough groceries for? | ☐One Day ☐3 Days ☐One Week ☐2 Weeks<br>☐3 Weeks ☐One Month ☐More than one month |
| P14. How many times did you go out last week? |  |
| P15. How many times did you visit someone's home last week? |  |
| P16. How many people visited you last week? |  |
| P17. Do you stay at home during the time of the national sanitization program (8:00 pm to 6:00 am)? | ☐Yes ☐No ☐Sometimes |
| P18. How many meters are considered a safe social distance between you and others? |  |
| **Questionnaire of attitude towards SARS-COV-2 INFECTION** |  |
| Questions | Answer options |
| 1. What is your resource of Corona (SARS-COV-2 INFECTION) information? (Choose all that apply) | Newspaper; TV; radio news; social media including Twitter, Instagram, or Whatsapp; official websites including WHO and CDC webpages; other people; other resources. |
| 2. Which of the following is the most reliable source in your opinion? (Choose all that apply) | Newspaper; TV; radio news; social media including Twitter, Instagram, or Whatsapp; |
|  | official websites including WHO and CDC webpages; other people; other resources. |
| 3.Do you agree that Corona (SARS-COV-2 INFECTION) infection will finally be successfully controlled? | Yes, no, somehow, I don't know<br>If yes when? 1 week, 2 weeks ,1 month, 2-3 months, 4-6 months, >6 months |
| 4.If the vaccine arrives, I will be willing to get vaccinated. | ☐Yes ☐No ☐Not sure |

## Appendix 2. Risk assessment score

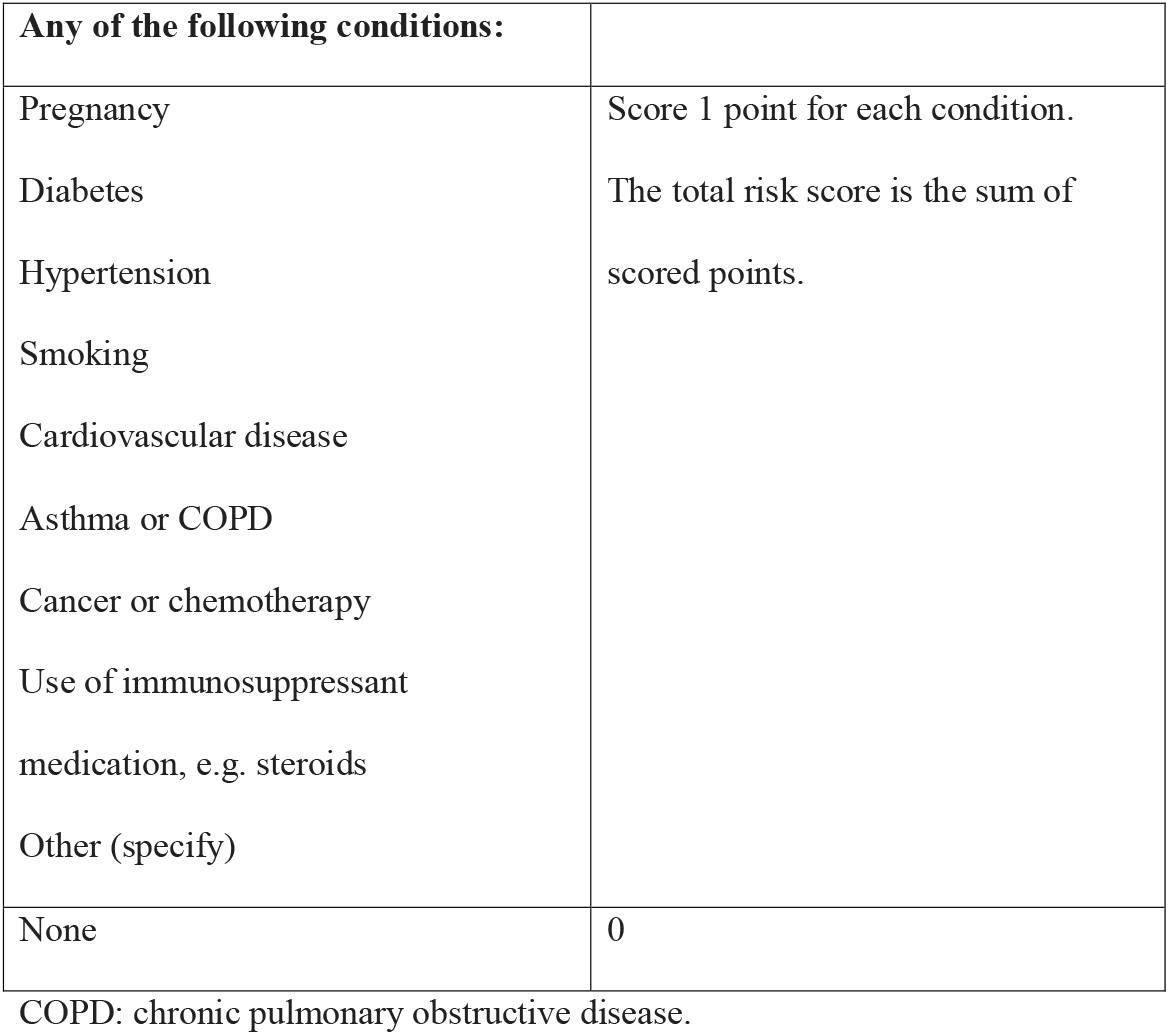

## Appendix 3. Patient Health Questionnaire-2 (PHQ-2)

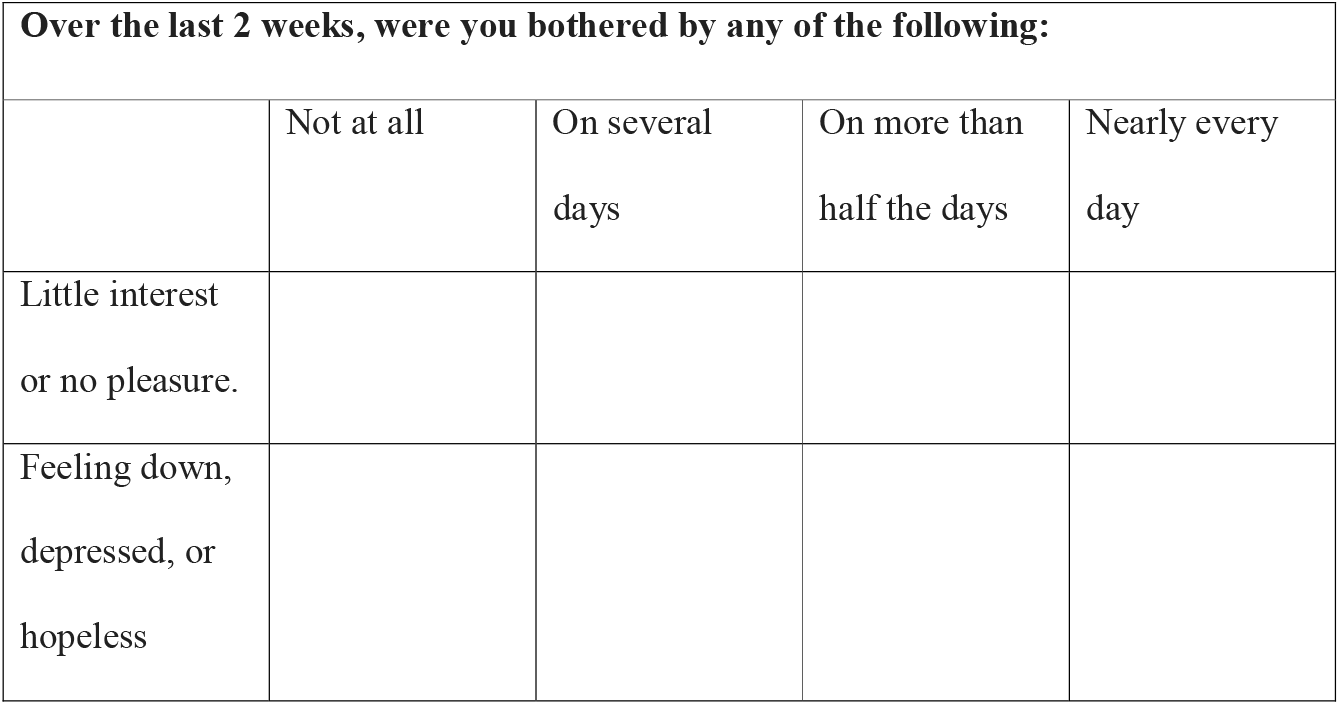

